# Combining Mass Spectrometry with Machine Learning to Identify Novel Protein Signatures: The Example of Multisystem Inflammatory Syndrome in Children

**DOI:** 10.1101/2025.04.17.25325767

**Authors:** Jeisac Guzmán Rivera, Haiyan Zheng, Benjamin Richlin, Christian Suarez, Sunanda Gaur, Elizabeth Ricciardi, Uzma N. Hasan, William Cuddy, Aalok R. Singh, Hulya Bukulmez, David C. Kaelber, Yukiko Kimura, Patrick W. Brady, Dawn Wahezi, Evin Rothschild, Saquib A. Lakhani, Katherine W Herbst, Alexander H Hogan, Juan C Salazar, Sandra Moroso- Fela, Jason Roy, Lawrence C. Kleinman, Daniel B. Horton, Dirk F. Moore, Maria Laura Gennaro

## Abstract

**Objectives:** We demonstrate an approach that integrates biomarker analysis with machine learning to identify protein signatures, using the example of SARS-CoV-2-induced Multisystem Inflammatory Syndrome in Children (MIS-C).

**Methods:** We used plasma samples collected from subjects diagnosed with MIS-C and compared them first to controls with asymptomatic/mild SARS-CoV-2 infection and then to controls with pneumonia or Kawasaki disease. We used mass spectrometry to identify proteins. Support vector machine (SVM) algorithm-based classification schemes were used to analyze protein pathways. We assessed diagnostic accuracy using internal and external cross-validation.

**Results:** Proteomic analysis of a training dataset containing MIS-C (N=17), and asymptomatic/mild SARS-CoV-2 infected control samples (N=20) identified 643 proteins, of which 101 were differentially expressed. Plasma proteins associated with inflammation and coagulation increased and those associated with lipid metabolism decreased in MIS-C relative to controls. The SVM machine learning algorithm identified a three-protein model (ORM1, AZGP1, SERPINA3) that achieved 90.0% specificity, 88.2% sensitivity, and 93.5% area under the curve (AUC) distinguishing MIS-C from controls in the training set. Performance was retained in the validation dataset utilizing MIS-C (N=17) and asymptomatic/mild SARS-CoV-2 infected control samples (N=10) (90.0% specificity, 84.2% sensitivity, 87.4% AUC). We next replicated our approach to compare MIS-C with similarly presenting syndromes, such as pneumonia (N=17) and Kawasaki Disease (N=13) and found a distinct three-protein signature (VWF, SERPINA3, and FCGBP) that accurately distinguished MIS-C from the other conditions (97.5% specificity, 89.5% sensitivity, 95.6% AUC). We also developed a software tool that may be used to evaluate other protein pathway signatures using our data.

**Conclusions:** We used MIS-C, a novel hyperinflammatory illness, to demonstrate that the use of mass spectrometry to identify candidate plasma proteins followed by machine learning, specifically SVM, is an efficient strategy for identifying and evaluating biomarker signatures for disease classification.

## Introduction

Timely diagnosis enables the timely delivery of effective treatment. When a new condition such as the Multisystem Inflammatory Disease in Children (MIS-C) emerges, researchers and clinicians seek both accurate paths to rapid diagnosis and effective treatments. MIS-C was first identified as an hyperinflammatory syndrome, representing a constellation of similar findings in the absence of an alternative explanation during SARS-CoV2 pandemic. Scientific teams worked to elucidate the linking pathophysiology, to establish paths to timely diagnosis, and to develop effective treatments. The field is still seeking accurate diagnosis and differentiation of MIS-C from other hyperinflammatory syndromes such as Kawasaki Disease (KD). We recently developed a proteomics analysis method which can be used as a diagnostic test for MIS-C. One of the dramatic consequences of infection in children with severe acute respiratory syndrome coronavirus 2 (SARS-CoV-2) is MIS-C (1), a rare, severe, and at times fatal condition characterized by fever, systemic hyperinflammation, and multi-organ dysfunction which can develop 2-6 weeks after a SARS-CoV-2 infection (2). As with many emerging conditions, MIS-C is a diagnosis of exclusion, and it requires a multi-tiered search for an alternative explanation before diagnosis is established (3), which has proven expensive in terms of resources and time to diagnosis.

Several studies have investigated the landscape of plasma proteins in MIS-C to gain insight in its pathogenesis and identify biomarkers that are distinctive for MIS-C (4–6). Most were structured toward candidate protein discovery rather than assessing their ability to discriminate MIS-C from comparator diseases. In the present study, we integrated data-independent acquisition mass spectrometry (DIA-MS) (7), and artificial intelligence to develop an analytical framework for biomarker selection and validation. We used mass spectrometry to identify proteins; support vector machine (SVM) (8), a machine learning approach, to identify proteins distinguishing subjects as having MIS-C or an identified alternative disease; and receiver operating characteristics (ROC) curves to assess the resulting model’s discrimination accuracy. Our work also resulted in an open-access SVM-based analytical tool and a robust dataset that enable the validation of protein biomarker signatures for MIS-C.

## Materials and Methods

### Study recruitment

We enrolled participants ≤ 21 years old (**Table 1**) and collected blood samples at nine sites from four states (CT, NJ, NY, OH). Children and youth with MIS-C were classified in accordance with the 2020 U.S. Centers for Disease Control criteria, which include recent history of SARS-CoV-2 infection, signs of inflammation and involvement of at least two organ systems, and no alternative plausible diagnosis (3). Pneumonia was defined by the presence of an infiltrative process in the lung parenchyma on chest radiography secondary to infection (viral or bacterial), without evidence of concurrent SARS-CoV-2 infection. Diagnosis of Kawasaki disease (KD) was based on established criteria (9). All pneumonia and KD participants tested negative for SARS-CoV-2 at enrollment. For all disease conditions, blood for proteomics analysis was collected during hospitalization. Controls in our study were subjects with a history of mild or asymptomatic SARS-CoV-2 infection were defined as having a positive SARS-CoV-2 test and presenting in the outpatient setting with no symptoms or symptoms not requiring inpatient care prior to sample collection.

**Table 1.** Study demographics of the combined training and validation data sets.

| **Demographics** | **MIS-C (n = 34)** | **Recovered**<br>**mild/asymptomatic**<br>**SARS-CoV-2 (n = 30)** | **Pneumonia (n = 17)** | **Kawasaki Disease**<br>**(n = 13)** |
| --- | --- | --- | --- | --- |
| **<u>Age (years)</u>** |  |  |  |  |
| 0-11 | 23 (63.9%) | 19 (63.3%) | 13 (76.5%) | 9 (69.2%) |
| 12-21 | 13 (36.1%) | 11 (36.7%) | 4 (23.5%) | — |
| Unknown | — | — | — | 4 (30.8%) |
| **<u>Sex</u>** |  |  |  |  |
| Female | 16 (44.4%) | 16 (53.3%) | 5 (29.4%) | 5 (38.5%) |
| Male | 20 (55.6%) | 14 (46.7%) | 12 (70.6%) | 8 (61.5%) |
| **<u>Race / Ethnicity</u>** |  |  |  |  |
| American Indian | — | — | — | 1 (7.7%) |
| Hispanic | 14 (38.9%) | 1 (3.3%) | — | 6 (46.2%) |
| Non-Hispanic Asian | 1 (2.8%) | 4 (13.3%) | 2 (11.8%) | 2 (15.4%) |
| Non-Hispanic Black | 3 (8.3%) | 5 (16.7%) | 2 (11.8%) | — |
| Non-Hispanic White | 10 (27.8%) | 20 (66.7%) | 12 (70.6%) | 3 (23%) |
| Unknown | 5 (13.9%) | — | 1 (5.8%) | 1 (7.7%) |

### Study approval

All study activities were approved by the Rutgers Institutional Review Board (Pro2020002961) and all participants provided informed consent prior to engaging in study activities.

### Sample preparation for mass spectrometry

Plasma (10µg per sample) was diluted in 50 mM HEPES, 50 mM EDTA and 2% in SDS and reduced with 5mM DTT for 30 minutes at 60°C and alkylated with 20mM iodoacetamide for 1 hour at room temperature in the dark. The sample was then subjected to SP3 beads digestion with trypsin (sequencing grade, Thermo Scientific) in 100mM ammonium bicarbonate, 2mM CaCl2 and incubated at 37°C overnight, as described (10). Peptides were acidified with formic acid and 10.0% of each sample was analyzed by liquid chromatography-tandem mass spectrometry (LC-MS/MS).

### Liquid chromatography-tandem mass spectrometry (LC-MS/MS)

Samples were analyzed by data-independent acquisition mass-spectrometry (7) using a Dionex Ultimate 3000 RLSCnano System (Thermo Fisher Scientific) interfaced with an Orbitrap Eclipse Tribrid mass spectrometer (Thermo Fisher Scientific). Raw data were analyzed using an in-silico predicted peptide library generated from the UniProt human reference proteome for library-free database searching using DIA-NN 1.8.1 (11). Results were filtered for posterior error probability (PEP) for the precursor identification of <1% and Protein Group Q value, also < 1%. Protein abundance was expressed as protein group MaxLFQ values (12).

### Data analysis methods

Study participants were divided into (i) a training set comprising 20 control participants with a history of mild or asymptomatic SARS-CoV-2 infection, and 17 participants with MIS-C and (ii) a validation set comprising 10 control mild/asymptomatic participants, 17 MIS-C participants, 17 pneumonia participants, and 13 KD participants. From the proteins from which DIA MaxLFQ abundance values were generated, we excluded from analysis proteins whose values were below the limit of detection in at least 50% of the samples. MaxLFQ values were log2 transformed prior to statistical analysis. We fitted a linear model comparing diseased to control for each protein. The result of these fitted models was, for each protein, an estimate of the log2 abundance ratio and its standard error, from which we calculated a p-value. To adjust for multiple comparisons, we converted the raw p-values to Holm p-values (13). We also calculated q-values from the raw p-values using the Benjamini-Hochberg method.

### Classification model building

We used the SVM classifier (R function “svm” in the “e1071” package) to develop models using the DIA-MS protein data to distinguish MIS-C from other conditions. We calculated sensitivity, specificity, and area under the ROC curve (AUC) (14) to assess the accuracy of SVM models. We next built a classifier model based on the currently available set of patients (the “training” set) and applied it to an external set of patients (the “validation” set) to obtain an externally validated AUC. Five random repetitions of five-fold cross-validation were used to calculate 95% confidence intervals for the AUC. We also developed an R package “miscClassify” that allows researchers to input candidate protein signatures and determine their performance with our validation data set. The supplemental material describes how to install and use the package, which is available on GitHub (https://github.com/mooredf22/miscPredict/).

### Term enrichment analysis

Pathway analysis was conducted on a protein list derived from differential expression analysis using the R package Enrichr (version 3.2) (15). Enrichment analysis was performed using the Reactome and Gene Ontology (GO) Biological Processes databases. Each protein set enrichment was assessed by Fisher’s Exact Test, and results were filtered by requiring a false discovery rate (FDR) < 0.05.

## Results

### The MIS-C plasma proteome

Proteomic analysis was conducted with the training set comprising 17 MIS-C and 20 mild/asymptomatic SARS-CoV-2 infection control samples. A total of 1,675 proteins were identified by DIA-MS. After removing the proteins with >50% missing values, we retained 643 proteins. Of these, 101 were found to be differentially abundant between the MIS-C and control groups based on Benjamini-Hochberg adjusted q-value (q<0.05), which corresponds to an FDR of 5 percent. Of the 101 differentially expressed proteins, 41 were more abundant and 60 were less abundant in MIS-C than in control samples (**Figure 1**). We performed gene ontology and pathway enrichment analysis and found that the top 20 enriched terms for the differentially increased proteins included terms related to immune function (**Figure 2A** and **2B**). The top 20 enriched terms for the differentially decreased proteins include lipid metabolism, coagulation, and protein metabolism (**Figure 2C** and **2D**). These findings emphasize the involvement of immune dysregulation, lipid metabolism, and coagulation pathways in its pathophysiology.

**Figure 1.**
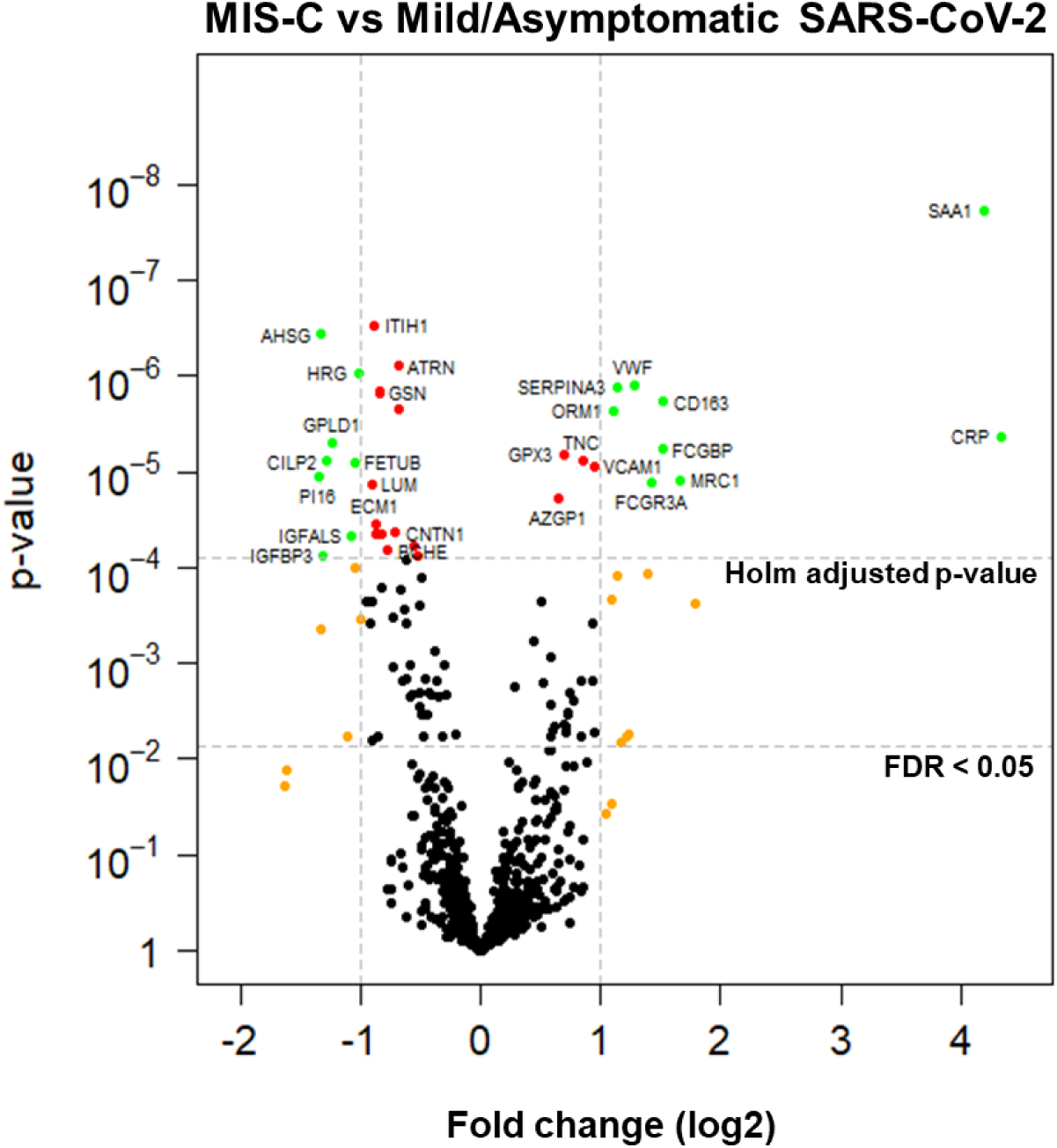
Volcano plot of differentially abundant proteins between MIS-C and mild/asymptomatic SARS-CoV-2. Green and red circles are proteins with a Holm’s adjusted p-value < 0.05. Green and orange circles are proteins with at least a 2-fold change.

**Figure 2.**
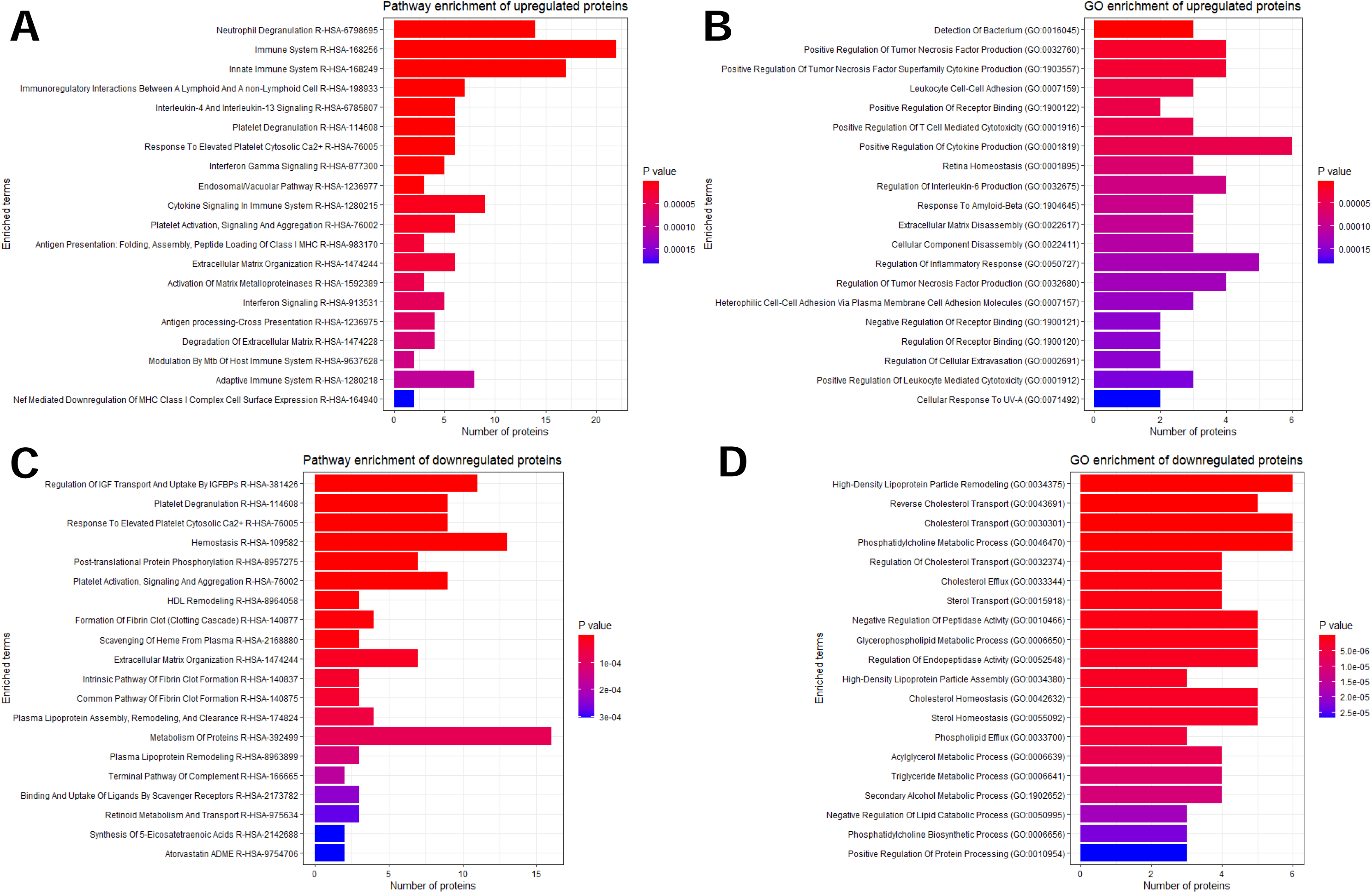
Term enrichment analysis of differentially abundant proteins. Proteins shown gave an FDR < 0.05 and an adjusted p-value < 0.05 in MIS-C vs. Mild/Asymptomatic SARS-CoV-2. Top 20 enriched A) Gene ontology and B) Reactome terms of differentially increased proteins in MIS-C. Top 20 enriched C) Gene ontology and B) Reactome terms of differentially decreased proteins in MIS-C.

### Development of a Support Vector Machine (SVM) model

To develop a plasma protein signature, we first used the Holm correction (13), which resulted in 34 proteins having a corrected *p*-value of ≤0.05. To evaluate the ability of these proteins to distinguish MIS-C from mild/asymptomatic cases we employed an SVM machine-learning algorithm. We selected proteins using three criteria: i) Holm corrected p-value, ii) intercept, a coefficient that accounts for protein abundance levels, and iii) increased abundance in MIS-C relative to controls. The latter criterion was applied since biomarker level increase may be suitable for the downstream development of immunodiagnostic assays for clinical use. We used the top three proteins (ORM1, SERPINA3, AZGP1) (Table 2) to build an SVM classifier model. This model exhibited high specificity (90.0%) and sensitivity (88.2%), and an area-under-the-curve (AUC) of 93.5% (CI 84.8% – 100%). Using two proteins yielded a lower AUC (90.4%; CI 86.8% – 94.0%), while adding more proteins to the model did not improve its characteristics. We next performed external validation of the model by utilizing the validation set of 17 MIS-C and 10 mild/asymptomatic infection control samples to match the conditions used in the training set. The resulting model showed a specificity of 90.0%, a sensitivity of 84.2%, and an AUC of 87.4% (CI 74.1% – 100%) (**Figure 3** and **Table 3**), showing that our SVM algorithm can predict MIS-C with high accuracy.

**Table 2.** Protein candidates used to build the predictive model.

| **Protein** | **Intercept** | **Fold change (log2)** | **Holm adjusted p-value** |
| --- | --- | --- | --- |
| **ORM1** | 27.05 | 1.10 | 1.47E-03 |
| **SERPINA3** | 26.58 | 1.14 | 8.28E-04 |
| **AZGP1** | 23.83 | 0.65 | 1.16E-02 |
| **GPX3** | 21.86 | 0.70 | 4.20E-03 |
| **FCGBP** | 20.26 | 1.51 | 3.69E-03 |
| **VWF** | 19.94 | 1.29 | 7.96E-04 |
| **CRP** | 19.00 | 4.34 | 2.67E-03 |
| **VCAM1** | 17.96 | 0.94 | 5.55E-03 |
| **TNC** | 17.87 | 0.85 | 4.74E-03 |
| **SAA1** | 17.58 | 4.19 | 1.22E-05 |
| **CD163** | 17.55 | 1.52 | 1.18E-03 |
| **FCG3RA** | 16.42 | 1.43 | 8.06E-03 |
| **MRC1** | 16.20 | 1.66 | 7.57E-03 |

Proteins are ordered by p-value and only proteins upregulated in MIS-C are shown. The intercept is a coefficient that represents the mean of the log2 transformed protein abundance of the control samples.

**Figure 3.**
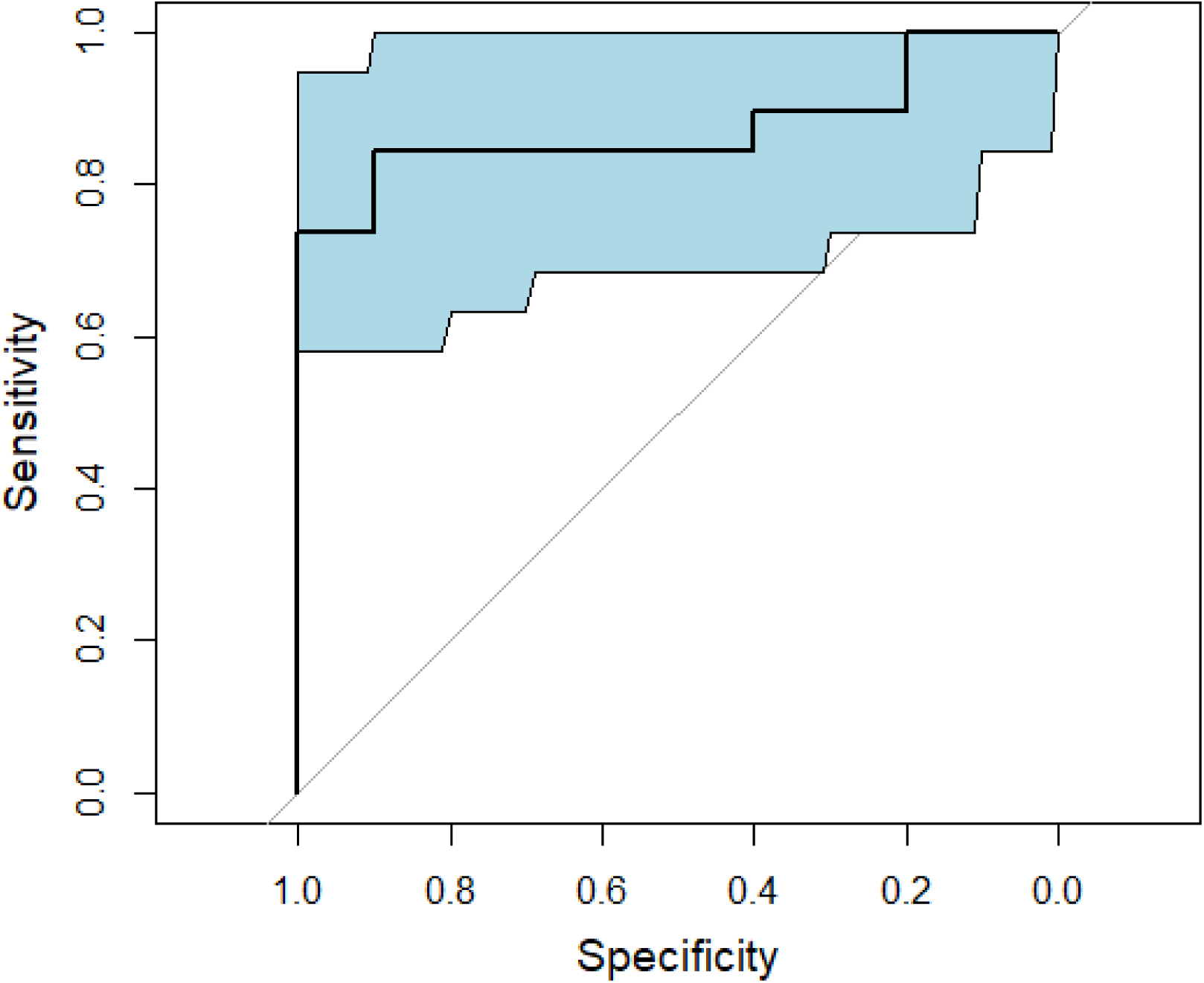
External validation of an SVM model. The figure shows a receiver operating characteristic (ROC) curve visualizing the performance of three proteins (ORM1, SERPINA3, and AZGP1) applied to the validation dataset (MIS-C vs. Mild/Asymptomatic SARS-CoV-2).

**Table 3.** Support Vector Machine model evaluation.

|  | **Overall accuracy** | **Sensitivity** | **Specificity** | **AUC** |
| --- | --- | --- | --- | --- |
| **Training set** | 89.2% | 88.2% | 90.0% | 93.5% |
| **Validation set** | 86.2% | 84.2% | 90.0% | 87.4% |

The values in the training set present error estimates which are not corrected for overfitting. The validation set values present error rate estimates that are calculated on the external validation set. AUC: area-under-the-curve

### Identification and validation of a multi-protein signature of MIS-C

In the clinical setting, it is necessary to distinguish MIS-C from other pathologies presenting with similar signs and symptoms. Therefore, we performed DIA-MS on samples obtained from pneumonia and KD patients to identify protein biomarkers that can accurately differentiate these conditions from MIS-C. We first performed pairwise comparisons between MIS-C and each comparator condition to identify distinct protein expression patterns (**Figures 4A-4C**). We observed that von Willebrand factor (VWF) was significantly increased in MIS-C on all comparisons, and this was the only protein that reached statistical significance in the comparison between MIS-C and KD (**Figures 4A** and **4D**). We also observed that the proteins FCGBP, VWF, F11, BCHE, KLKB1, ATRN, SERPINA3, A2M, and PGLYRP2 were shared when comparing MIS-C against pneumonia and mild/asymptomatic SARS-CoV-2 infection (**Figures 4B-4D**). When we compared MIS-C to all groups in a multi-disease comparison we observed that FCGBP, VWF, and SERPINA3 were the top three upregulated proteins in MIS-C, out of 33 proteins that had a Holm p-value ≤ 0.05 (**Figure 5A** and **Table 4**). An SVM model utilizing these three proteins showed a sensitivity of 89.5%, specificity of 97.5%, and an AUC of 95.6% (CI 89.6% – 100%) (**Figure 5B**) A two-protein model gave comparable results, while adding more proteins did not increase model’s accuracy. To correct our three-protein model for overfitting, we next carried out five-fold cross-validation using the “crossval” R library, and found a sensitivity of 75.3%, a specificity of 92.0%, and an AUC of 93.4% (CI 90.0% – 100%). While these cross-validated estimates are lower than the uncorrected ones, as expected, they remained high. These results indicate that a plasma protein set comprising VWF, SERPINA3, and FCGBP exhibits a strong predictive capability for distinguishing MIS-C from pneumonia, KD, and mild/asymptomatic cases in children.

**Figure 4.**
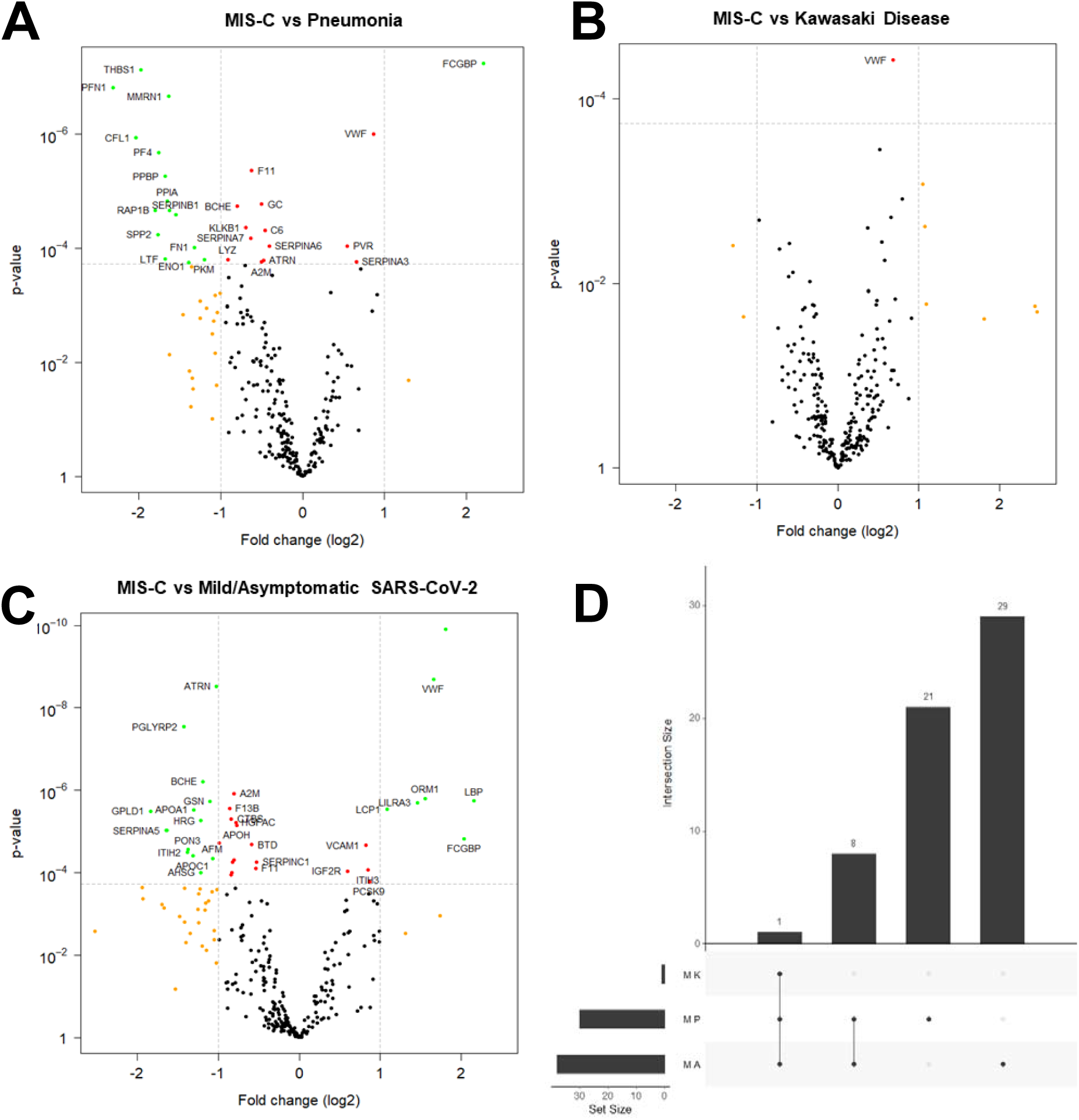
Differentially abundant proteins between MIS-C, pneumonia, Kawasaki Disease, and mild/asymptomatic SARS-CoV-2. Volcano plots of differentially abundant proteins between A) MIS-C and mild/asymptomatic SARS-CoV-2, B) MIS-C and pneumonia, and C) MIS-C and Kawasaki disease. Green and red circles are proteins with a Holm’s adjusted p-value < 0.05. Green and orange circles are proteins with at least a 2-fold change. D) UpSet plot showing the shared proteins between all pairwise comparisons. MK: MIS-C vs Kawasaki Disease; MP: MIS-C vs Pneumonia; MA: MIS-C vs Mild/Asymptomatic SARS-CoV-2

**Figure 5.**
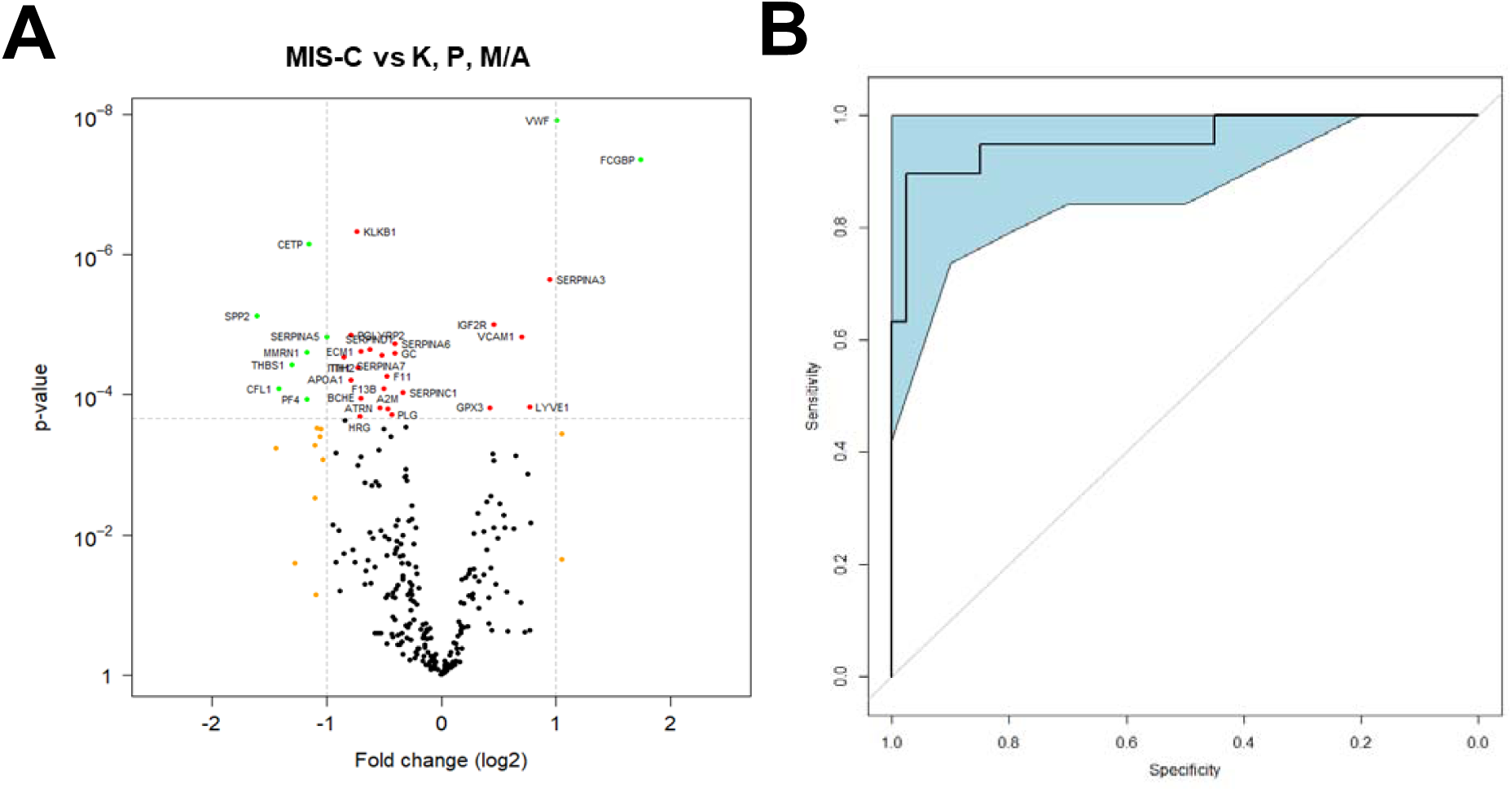
Multi-disease comparison and SVM model. A) Volcano plot of differentially abundant proteins between MIS-C and Kawasaki disease (K), pneumonia (P), and mild/asymptomatic SARS-CoV-2 (M/A). Green and red circles are proteins with a Holm’s adjusted p-value < 0.05. Green and orange circles are proteins with at least a 2-fold change. B) Receiver operating characteristic (ROC) curve visualizing the performance of a 3-protein signature (VWF, FCGBP, and SERPINA3). K: Kawasaki disease; P: Pneumonia; M/A: Mild/Asymptomatic SARS-CoV-2

**Table 4.** Biomarker candidates from multi-disease comparison.

| **Protein** | **Fold change (log2)** | **Holm adjusted p-value** |
| --- | --- | --- |
| **FCGBP** | 1.74 | 1.19E-05 |
| **VWF** | 1.01 | 3.29E-06 |
| **SERPINA3** | 0.95 | 6.07E-04 |
| **LYVE1** | 0.77 | 3.66E-02 |
| **VCAM1** | 0.70 | 3.84E-03 |
| **IGF2R** | 0.46 | 2.61E-03 |
| **GPX3** | 0.43 | 3.67E-02 |

Proteins are ordered by p-value and only proteins upregulated in MIS-C are shown.

### Model validation using external protein markers

Multiple groups have studied the plasma proteome of MIS-C patients for biomarker discovery. However, very few have applied classification models to their differentially expressed proteins. To address this gap, we applied the analytical tool described in this work to independently evaluate the performance of two multi-protein signatures for which an AUC was calculated in recent publications (**Table 5**). Nygaard et al. (2024) proposed a signature of “FCGR3A”, “LCP1”, “SERPINA3”, “BCHE”, distinguishing MIS-C from KD and bacterial and viral infections. They reported an AUC of 95.0% based on internal validation and 87.0% on an external validation (16). When we used our dataset and SVM model to assess their biomarker panel, we achieved comparable AUC values of 95.8% (uncorrected) and 89.7% (cross-validated corrected estimate). Similarly, Yeoh et al. (2024) described a signature based on “CD163”, “PCSK9”, and “CXCL9”, which also distinguished MIS-C from KD and bacterial and viral infections, with an originally reported incorrected AUC of 85.7% (17). We built an SVM model with two of these proteins (chemokines such as CXCL9 are not detected by our DIA-MS method) and achieved an AUC of 94.1% (uncorrected) and 82.4% (cross-validated correction) (**Table 5**). These results show that the analysis pipeline and the proteomics dataset generated in the present study can be applied to the evaluation of the classification performance of differentially expressed proteins identified in independent studies of MIS-C.

**Table 5.** Comparison of model performance.

| Signature | Proteins | Reference | Original AUC | AUC and 95% Confidence Interval | CV-corrected AUC |
| --- | --- | --- | --- | --- | --- |
| 1 | ORM1, SERPINA3, AZGP1 | Current manuscript | ————— | 88.0%<br>(77.4% - 98.7%) | 79.7% |
| 2 | VWF, FCGBP, SERPINA3 | Current manuscript | ————— | 95.6%<br>(89.6% - 100%) | 94.2% |
| 3 | LCP1, SERPINA3, BCHE | Nygaard et al. 2024 | 100% (test set)<br>94% (internal validation)<br>87% (external validation) | 95.8%<br>(91.2% - 100%) | 89.7% |
| 4 | CD163, PCSK9, [CXCL9]* | Yeoh et al. 2024 | 85.7%<br>(76.6% - 94.8%) | 94.1%<br>(87.8% - 100%) | 82.4% |

\*Yeoh (2024) included CXCL9 in their protein signature, but we excluded it since we have no data for that protein. AUC: area-under-the-curve; CV: cross-validation.

## Discussion

We describe an empirical approach to identify biomarkers for the classification of inflammatory illnesses, using the example of MIS-C, a syndrome for which specific biomarkers accelerating the diagnostic process are still missing. By integrating clinical epidemiology, mass spectrometry, and SMV machine learning, we identified a small set of proteins for MIS-C classification. Our work also developed an SVM-based classification algorithm for refinement and validation of previously identified MIS-C biomarkers that were not externally validated.

Our analysis of the MIS-C plasma proteome contributes to the understanding of MIS-C pathogenesis and its underlying biological mechanisms. These include disruption of coagulation, which may worsen vascular damage and inflammation (18), and increased abundance of proteins associated with neutrophil degranulation, cytokine production, and immune response to pathogens is associated with hyperinflammation (19). We also observed reduced plasma levels of proteins involved in metabolism of cholesterol, phospholipids, triglycerides, and various lipoproteins, which have been previously associated with severity and unfavorable outcome of SARS-CoV-2 infection manifestations in children (20). Our plasma proteome analysis is consistent with previous reports (4, 6). Most importantly, the three-protein diagnostic biosignature we identified is fully consistent with MIS-C pathogenesis.

Plasma levels of SERPINA3, a member of the serine protease inhibitor superfamily, are increased during acute inflammation to control inflammation by prevention of diapedesis and phagocytosis by neutrophils to prevent tissue damage, therefore its plasma levels correlate with duration of the inflammatory phase and multiorgan damage (21, 22), which is a hallmark of MIS-C. There are observations and studies showing that SERPINA3 is elevated during viral infections such as human rhinovirus (23) and particularly during SARS-CoV2 infections in vivo (24) and in vitro (25). Elevated levels of SERPINA3 and in certain cancers, where they are associated with poor outcome (21).

Increased plasma levels of IgGFc-binding protein (FCGBP), a mucin-like protein that mediates transport of serum IgG to mucosal surfaces and contributes to mucosal immunity (26), can be explained by the damage of the gut mucosal barrier observed in MIS-C (27, 28). Elevated levels of von Willebrand factor (VWF), which promotes hemostasis and platelet adhesion, indicates endothelial activation and damage, leading to formation of microvascular thrombi and disseminated intravascular coagulation (18, 29), which are indicators of the hypercoagulable state observed in MIS-C (30). Thus, our results well connect biomarker discovery with pathogenesis.

Our study has limitations. The demographic and geographic variability of plasma protein levels may need further biosignature validation in diverse populations. Moreover, a definitive catalog of MIS-C biomarkers would benefit from expanding the control conditions to other hyperinflammatory syndromes (e.g., systemic juvenile idiopathic arthritis and hemophagocytic lymphohistiocytosis) and conditions affecting intestinal permeability, such as inflammatory bowel disease. Additionally, our data were collected during disease management, and we were unable to account for patient treatment, which might impact plasma protein levels. Moreover, our cross-sectional design did not include collection of longitudinal samples, precluding the temporal analysis of biomarker dynamics.

In conclusion, our approach, which integrated clinical epidemiology, mass spectrometry, and artificial intelligence, shifts the focus of MIS-C biomarker research from mere discovery to differential diagnosis. Further work will help expand the evaluation of our markers of MIS-C to more complex clinical settings and the application of our modeling tools to finding classification biomarkers for other challenging hyperinflammatory syndromes including KD, macrophage activation syndrome, and hemophagocytic lymphohistiocytosis. We also expect our work to contribute to preparedness to the potential resurgence of MIS-C and the advent of new syndromes.

## Data Availability

All data produced in the present study are available upon request to the authors.

## Acknowledgements

DIA mass spectrometry was performed by the Biological Mass Spectrometry Facility at Rutgers Robert Wood Johnson Medical School. We wish to thank David Sleat for his contribution in establishing and executing the DIA MS pipeline. This work was funded by NIH grants R61HD105619, R33HD105619, HD105593-03S2, R01AI158911, HD105613, and NCATS UM1TR004789, and by Rutgers ROI–HealthAdvance HA2022-0039.

